# Stringent thresholds for SARS-CoV-2 IgG assays result in under-detection of cases reporting loss of taste/smell

**DOI:** 10.1101/2020.07.21.20159038

**Authors:** David W Eyre, Sheila F Lumley, Denise O’Donnell, Nicole E Stoesser, Philippa C Matthews, Alison Howarth, Stephanie B Hatch, Brian D Marsden, Stuart Cox, Tim James, Richard J Cornall, David I Stuart, Gavin Screaton, Daniel Ebner, Derrick W Crook, Christopher P Conlon, Katie Jeffery, Timothy M Walker, Timothy EA Peto

**Author notes:** Corresponding author: David Eyre.

## Abstract

Thresholds for SARS-CoV-2 antibody assays have typically been determined using samples from symptomatic, often hospitalised, patients. Assay performance following mild/asymptomatic infection is unclear. We assessed IgG responses in asymptomatic healthcare workers with a high pre-test probability of Covid-19, e.g. 807/9292(8.9%) reported loss of smell/taste. The proportion reporting anosmia/ageusia increased at antibody titres below diagnostic thresholds for both an in-house ELISA and the Abbott Architect chemiluminescent microparticle immunoassay (CMIA): 424/903(47%) reported anosmia/ageusia with a positive ELISA, 59/387(13.2%) with high-negative titres, and 324/7943(4.1%) with low-negative results. Adjusting for the proportion of staff reporting anosmia/ageusia suggests the sensitivity of both assays is lower than previously reported: Oxford ELISA 90.8% (95%CI 86.1-92.1%) and Abbott CMIA 80.9% (77.5-84.3%). However, the sensitivity may be lower if some anosmia/ageusia in those with low-negative titres is Covid-19-associated. Samples from individuals with mild/asymptomatic infection should be included in SARS-CoV-2 immunoassay evaluations. Reporting equivocal SARS-CoV-2 antibody results should be considered.

## Introduction

Serological testing for antibodies to SARS-CoV-2, the virus that causes Covid-19, has been used to estimate the extent of Covid-19 exposure in national^1,2^ and regional populations, as well as in subgroups of interest including healthcare workers.^3,4^ Although the extent of protection against future infection afforded by antibodies is not yet clear, most vaccine candidates aim to induce neutralizing antibodies against the viral spike protein^5^ and antibody-based “immunity passports” have been proposed as a possible option to support personalised exit from lockdown restrictions.

One key element of deploying antibody testing is determining assay thresholds for confirming the detection of antibodies. Typically, this has been done based on collections of pre-pandemic sera (i.e. known negative samples) and samples from patients with PCR-confirmed Covid-19 (deemed highly likely to be antibody positive).^6,7^ Often, these ‘known positive’ sera have been derived from individuals who accessed diagnostic testing by RT-PCR based on the presence of clinical symptoms, with a bias towards those with severe enough symptoms to present to hospital where diagnostic testing was concentrated early in the pandemic. As a result, it is not clear how applicable antibody assay thresholds are to those with prior asymptomatic or mild infection which may not have met syndromic criteria for testing, such as presence of fever. Furthermore, even in symptomatic individuals, those with milder disease may have lower viral loads^8^ and therefore be more likely to be falsely PCR-negative and omitted from assay calibration cohorts.

We have recently undertaken SARS-CoV-2 serological testing in a large cohort of UK healthcare workers.^3^ In keeping with other researchers,^9–11^ we reported that loss of smell (anosmia) or taste (ageusia) were highly predictive of Covid-19, the presence of either was associated with an odds ratio (OR) for Covid-19 of 17.7 (95%CI 14.1-22.3) after adjustment for other symptoms.^3^ A number of healthcare workers tested were surprised to be given negative antibody results following clinical syndromes consistent with Covid-19, including healthcare workers who were household contacts of PCR-confirmed cases. Therefore, here we use the presence of anosmia/ageusia along with other risk factors for Covid-19 as probes for Covid-19 infection that was potentially undiagnosed by immunoassays by investigating their relationship with antibody titres either side of the assay threshold.

## Results

9454 hospital staff provided serum samples between 23^rd^ April and 8^th^ June 2020, together with complete associated complete symptom and risk factor survey data. Of these samples, 9292 were analysed using an in-house ELISA developed in Oxford (targeting trimeric spike protein) and are considered in subsequent analyses; 903 (9.7%) had IgG detected. 9113 of these samples were also analysed using the Abbott Architect chemiluminescent microparticle immunoassay (CMIA) targeting nucleoprotein; 782 (8.6%) had IgG detected by both platforms, 113 (1.2%) by only Abbott and 103 (1.1%) by the Oxford ELISA only.

### Antibody titres in individuals with a prior PCR-positive nasopharyngeal swab

168 staff participating in asymptomatic testing had previously tested PCR-positive for SARS-CoV-2 on a nasopharyngeal swab, obtained as part of testing offered to symptomatic staff,≥14 days prior to their blood being obtained for serology. 152/168 (90.5%, 95%CI 85.0-94.5%) were IgG positive using the Oxford ELISA when tested between 14 and 62 days after their PCR test (Figure 1A) and 152/162 (93.8%, 95%CI 88.9-97.0%) using the Abbott CMIA (Figure 1B). There was no clear relationship between negative antibody results and time since a positive PCR test, although there was some evidence that Abbott CMIA titres fell over time, but remained above the diagnostic threshold, which was not seen with the Oxford ELISA which has more limited dynamic range above the diagnostic threshold.

**Figure 1.**
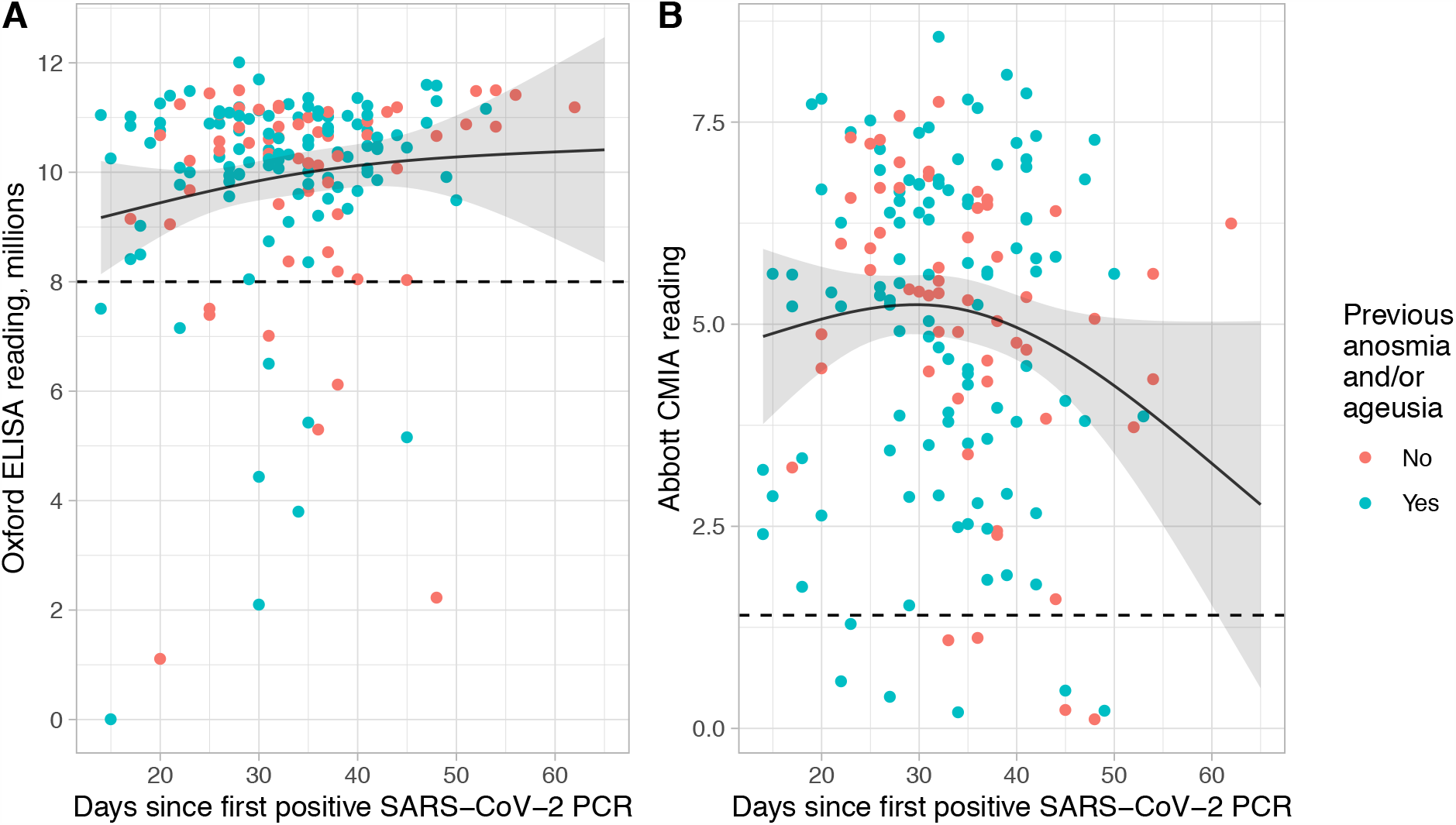
SARS-CoV-2 IgG antibody titres in convalescent symptomatic healthcare workers ≥14 days following a positive PCR test. Panel A shows titres using the Oxford ELISA assay targeting trimeric spike protein (n=168) and panel B shows titres using the Abbott CMIA targeting nucleocapsid protein (n=162). The dashed horizontal lines show the pre-defined threshold for reporting antibody detection (Oxford ELISA 8 million, Abbott CMIA 1.4). The solid line and ribbon shows the fitted mean value and 95% confidence interval using a linear regression model with a 3 knot spline. Points are coloured by whether staff reported previous anosmia (loss of smell) and/or ageusia (loss of taste) since 01 February 2020 when asked prior to serological testing.

### Predictors of anosmia and ageusia in seropositive healthcare workers

As only a minority of healthcare workers studied had a prior positive PCR test (168/9292, 1.8%), and these individuals had all been sufficiently symptomatic to access the test (fever≥37.8°C or a new persistent cough), we therefore investigated other less stringent clinical markers of possible Covid-19. Of the 903 staff with SARS-CoV-2 IgG antibodies detected using the Oxford ELISA, 424 reported loss of smell and/or taste since 01 February 2020 when asked before serum sampling. Within these 903 staff, there was no association between self-reported ethnicity or the speciality area staff worked in and anosmia/ageusia (Table 1). On univariable analysis there was some evidence that junior doctors and laboratory staff with SARS-CoV-2 IgG antibodies were more likely to report loss of smell/taste (OR 1.99 [95%CI 1.06-3.75; p=0.03] and 3.11 [1.06-9.09; p=0.04] respectively). In a multivariable model, male gender was associated with fewer reports of loss of smell/taste (0.67 [0.50-0.89; p=0.007]) as was each 10-year increase in age (0.88 [0.79-0.99; p=0.03).

**Table 1.** **Association between demographic and workplace factors and self-reported anosmia or ageusia in 903 healthcare workers with positive SARS-CoV-2 serology**.

|  |  | Total | Number reporting anosmia | Percentage with Covid-19 reporting anosmia/ ageusia | Univariable odds ratio for anosmia/ ageusia | 95% CI | p value | Multivariable odds ratio for anosmia | 95% CI | p value |
| --- | --- | --- | --- | --- | --- | --- | --- | --- | --- | --- |
| Ethnicity | White | 534 | 254 | 48% | 1.00 |  |  |  |  |  |
|  | Asian | 238 | 112 | 47% | 0.98 | (0.72, 1.33) | 0.90 |  |  |  |
|  | Black | 58 | 24 | 41% | 0.78 | (0.45, 1.35) | 0.37 |  |  |  |
|  | Chinese | 5 | 2 | 40% | 0.73 | (0.12, 4.43) | 0.74 |  |  |  |
|  | Mixed | 23 | 10 | 43% | 0.85 | (0.37, 1.97) | 0.70 |  |  |  |
|  | Not stated | 13 | 7 | 54% | 1.29 | (0.43, 3.88) | 0.66 |  |  |  |
|  | Other | 32 | 15 | 47% | 0.97 | (0.48, 1.99) | 0.94 |  |  |  |
| Role | Administrative staff | 73 | 30 | 41% | 1.00 |  |  |  |  |  |
|  | Biomedical scientist and laboratory staff | 19 | 13 | 68% | 3.11 | (1.06, 9.09) | 0.04 |  |  |  |
|  | Senior doctor | 45 | 16 | 36% | 0.79 | (0.37, 1.7) | 0.55 |  |  |  |
|  | Junior doctor | 86 | 50 | 58% | 1.99 | (1.06, 3.75) | 0.03 |  |  |  |
|  | Nurse, healthcare assistant | 455 | 220 | 48% | 1.34 | (0.81, 2.21) | 0.25 |  |  |  |
|  | Other | 84 | 36 | 43% | 1.07 | (0.57, 2.03) | 0.82 |  |  |  |
|  | Other allied health professional | 39 | 22 | 56% | 1.85 | (0.85, 4.07) | 0.12 |  |  |  |
|  | Porter, Domestic | 51 | 14 | 27% | 0.54 | (0.25, 1.17) | 0.12 |  |  |  |
|  | Physiotherapists, Occupational therapists, Speech and language therapists | 31 | 15 | 48% | 1.34 | (0.58, 3.13) | 0.49 |  |  |  |
|  | Security, Estates, Catering | 20 | 8 | 40% | 0.96 | (0.35, 2.62) | 0.93 |  |  |  |
| Speciality area | Other | 312 | 134 | 43% | 1.00 |  |  |  |  |  |
|  | Anaesthetics | 14 | 6 | 43% | 1.00 | (0.34, 2.94) | 1.00 |  |  |  |
|  | Emergency Medicine | 34 | 16 | 47% | 1.18 | (0.58, 2.4) | 0.65 |  |  |  |
|  | General Surgery, Urology, Plastics, Vascular, Cardiothoracic surgery | 46 | 22 | 48% | 1.22 | (0.65, 2.26) | 0.53 |  |  |  |
|  | Haematology, Oncology | 44 | 21 | 48% | 1.21 | (0.64, 2.28) | 0.55 |  |  |  |
|  | Infectious Diseases, Respiratory | 21 | 8 | 38% | 0.82 | (0.33, 2.03) | 0.66 |  |  |  |
|  | Intensive Care Medicine | 37 | 16 | 43% | 1.01 | (0.51, 2.01) | 0.97 |  |  |  |
|  | Medicine | 179 | 92 | 51% | 1.40 | (0.97, 2.03) | 0.07 |  |  |  |
|  | Obstetrics and Gynaecology | 13 | 5 | 38% | 0.83 | (0.27, 2.59) | 0.75 |  |  |  |
|  | Ophthalmology, Ear, Nose and Throat, Maxillofacial surgery | 8 | 7 | 88% | 9.30 | (1.13, 76.48) | 0.04 |  |  |  |
|  | Paediatrics | 37 | 25 | 68% | 2.77 | (1.34, 5.71) | 0.01 |  |  |  |
|  | Radiology | 29 | 16 | 55% | 1.63 | (0.76, 3.51) | 0.21 |  |  |  |
|  | Specialist Medicine | 88 | 37 | 42% | 0.96 | (0.6, 1.56) | 0.88 |  |  |  |
|  | Trauma and Orthopaedics, Rheumatology | 41 | 19 | 46% | 1.15 | (0.6, 2.21) | 0.68 |  |  |  |
| Gender | Female | 637 | 319 | 50% |  |  |  |  |  |  |
|  | Male | 263 | 104 | 40% | 0.65 | (0.49, 0.87) | 0.004 | 0.67 | (0.50, 0.89) | 0.007 |
|  | Prefer not to say | 2 | 1 | 50% | 1.00 | (0.06, 16.01) | 1.00 | 1.17 | (0.07, 18.88) | 0.91 |
|  | Trans | 1 | 0 | 0% | 0.00 | (0.00, >100) | 0.97 | 0.00 | (0.00, >100) | 0.97 |
| Age | per 10 year increase |  |  |  | 0.87 | (0.78, 0.98) | 0.02 | 0.88 | (0.79, 0.99) | 0.03 |

### Relationship between anosmia/ageusia and antibody titres

A total of 807/9292 (8.9%) staff reported loss of smell/taste. In those with anosmia/ageusia who could recall a date of symptom onset, the large majority, 630/636 (99%), underwent serum sampling for serology ≥14 days after their symptom onset; a median (IQR) 45 (35-56) days later. We did not ask staff about duration of anosmia/ageusia. The 6 staff sampled earlier than 14 days post-symptom onset included 1 individual with IgG detected by both assays, 1 with an equivocal Oxford ELISA result and negative Abbott CMIA result, the remaining 4 had low negative results by both assays.

The proportion of staff reporting loss of smell/taste increased at antibody titres below the diagnostic threshold for both the Oxford ELISA (Figure 2A) and Abbott CMIA (Figure 2B), consistent with possible Covid-19 in those classified as having high negative (equivocal) antibody titres. The diagnostic threshold for detection of antibody by the Oxford ELISA is 8 million. However, from titres of 4 million and above the proportion of staff reporting loss of smell/taste started to rise from a baseline of ∼4% to 30% at titres of 7 million (0-3 million vs. 4 million: exact p=0.14 and vs. 5 million: p=0.008). Similarly, Abbott CMIA readings of 0.2 and above (diagnostic threshold 1.4) were associated with increased loss of smell/taste (<0.2 vs. 0.2-0.39: p<0.001). Similar trends were seen for antibody titres and self-reported fever and myalgia (Supplementary Figures S1 and S2).

**Figure 2.**
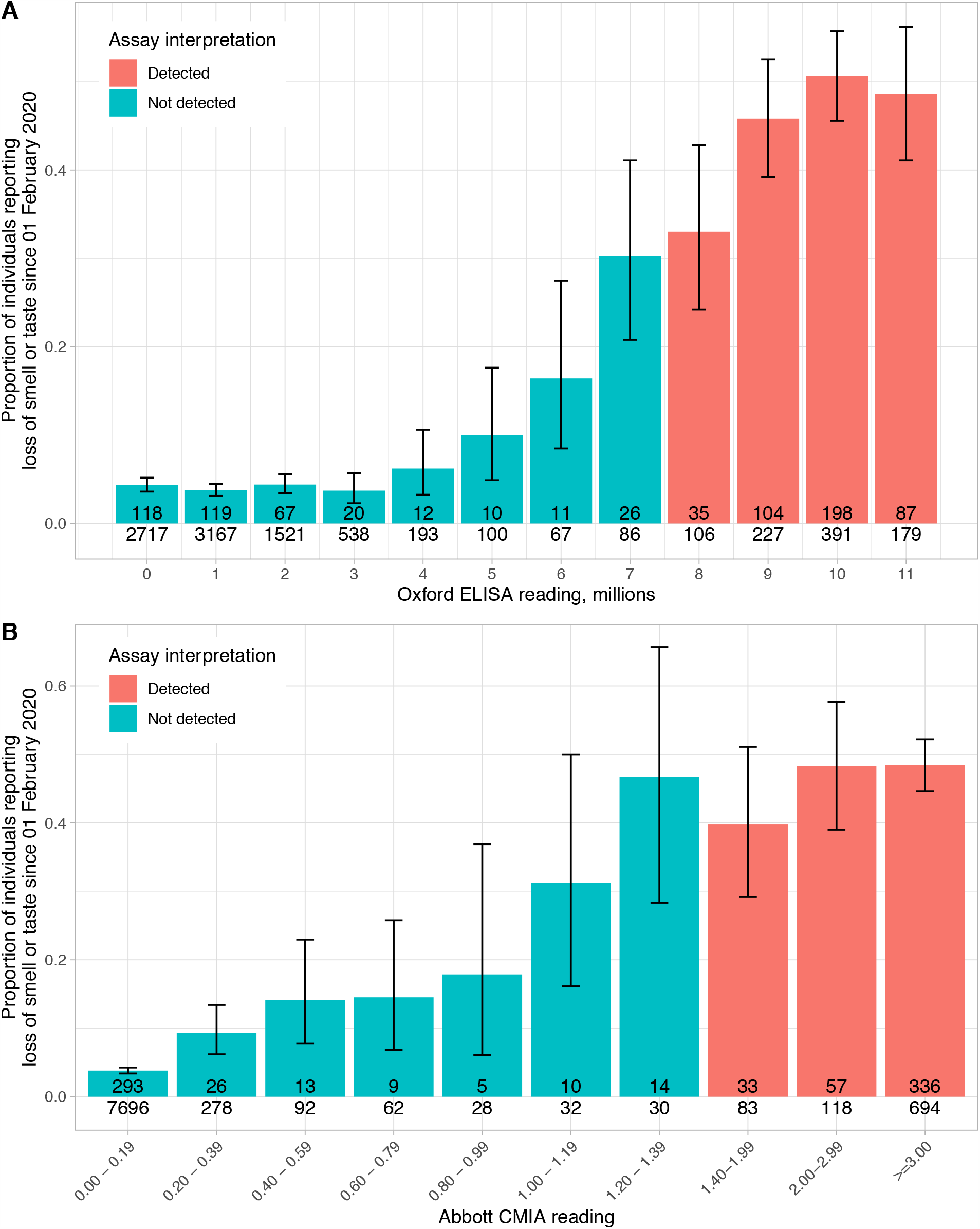
Proportion of staff with anosmia or loss of taste by antibody titre. Panel A shows the results using a trimeric spike ELISA and panel B the results from the Abbott CMIA targeting nucleocapsid protein, with blue showing results called negative and red showing those called as positive based on pre-defined assay thresholds. The number of individuals with these symptoms is shown in each bar, and the total number of individuals with each antibody titre below the bar. The error bars show 95% confidence intervals. For the Oxford ELISA readings each value is rounded down, such that for example a value of 1.7 million is within the 1 million bar.

Regarding Oxford ELISA readings between 4 and 8 million and Abbott CMIA readings between 0.2 and 1.4 as equivocal, equivocal results by one assay were frequently associated with positive results by the other (Figure 3A). However, we also observed an increase in anosmia/ageusia in staff with equivocal Abbott readings where Oxford ELISA results were equivocal or low negative. However, the incidence of anosmia/ageusia was near baseline with low negative Abbott readings irrespective of the Oxford ELISA reading (Figure 3B).

**Figure 3.**
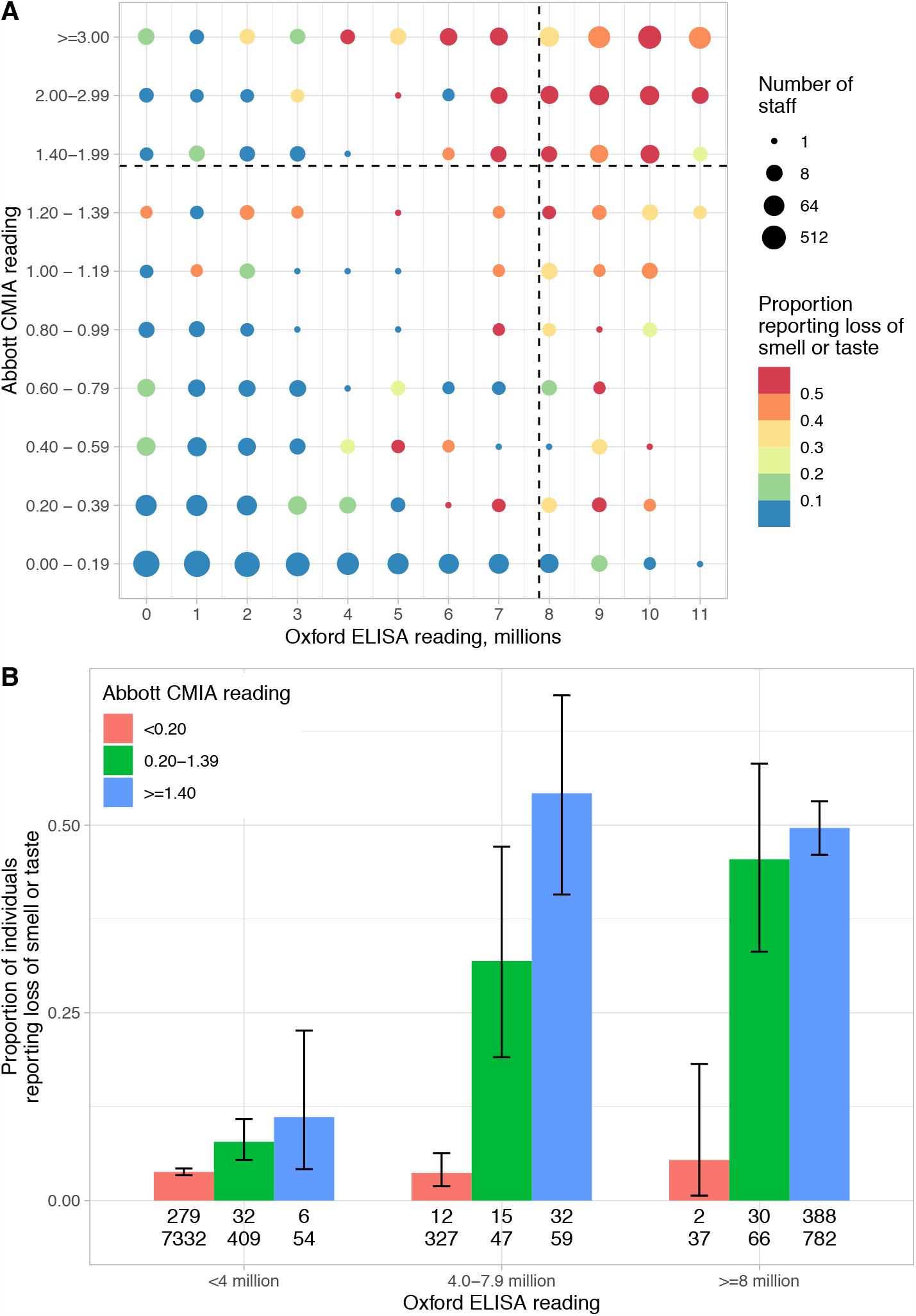
The relationship between Abbott CMIA and Oxford ELISA readings and loss of smell or taste. Panel A compares the number of individuals with combinations of Abbott CMIA and Oxford ELISA titres, the size of each circle represents the number of individuals and the colour the proportion reporting loss of smell or taste. Panel B groups the data by positive, high-negative (equivocal) and low-negative titres for both assays, the numbers shown beneath each bar are the number of individuals reporting loss of smell/taste and the total number of individuals with the antibody titre.

### Impact on estimated serology sensitivity for Covid-19

To estimate the extent to which current antibody thresholds may under-estimate past Covid-19 infections we considered the percentage of individuals reporting loss of smell/taste by 3 antibody titre categories (Table 2). Conservatively, considering those with an Oxford ELISA of <4 million reporting loss of smell/taste as the background rate of anosmia/ageusia unrelated to Covid-19, an estimated additional 9.1% of individuals with antibody titres of 4.0-7.9 million reported Covid-19-associated anosmia/ageusia, and 42.9% with titres of >8 million. Assuming that the sensitivity of antibody tests is similar in those reporting and not reporting loss of smell/taste, and taking the previously reported sensitivity of the Oxford ELISA as 99.1% (531/536),^7^ the true sensitivity may actually be 90.8% (95%CI 86.1-92.1%). However, more individuals with a previous positive PCR and low antibody titres reported anosmia than would be expected by a background rate of 4.1% (Figure 1). Therefore, with an alternative example assumption that the background rate of anosmia/ageusia is 3%, i.e. some of the anosmia/ageusia in those with low antibody titres is Covid-19-related, the sensitivity would be 75.1% (95%CI 67.6-81.4%). For Abbott the reported sensitivity of 92.7% (90.2-94.8%)^7^ was 80.9% (77.5-84.3%) after adjustment for a background rate of 3.8% (Table 2) and 70.8% (65.0-79.1%) assuming a 3% background rate of anosmia/ageusia.

**Table 2.** **Assessment of additional potential SARS-CoV-2 infections using loss of smell/taste and Oxford ELISA titres (n=9292 samples) and Abbott CMIA titres (n=9113)**.

|  | Oxford ELISA reading |  |  |
| --- | --- | --- | --- |
|  | <4 million<br>Low negative | 4.0-7.9 million<br>High negative | ≥8 million<br>Positive |
| Loss of smell/taste reported | 324 (4.1%) | 59 (13.2%) | 424 (47.0%) |
| Loss of smell/taste not reported | 7619 (95.9%) | 387 (86.8%) | 479 (53.0%) |
| Additional estimated number with loss of smell/taste above baseline | - | 35 (9.1%) | 387 (42.9%) |
| Total | 7943 | 387 | 903 |

|  | Abbott CMIA reading |  |  |
| --- | --- | --- | --- |
|  | <0.2<br>Low negative | 0.2-1.39<br>High negative | ≥1.4<br>Positive |
| Loss of smell/taste reported | 293 (3.8%) | 77 (14.8%) | 426 (47.6%) |
| Loss of smell/taste not reported | 7403 (96.2%) | 445 (85.2%) | 469 (52.4%) |
| Additional estimated number with loss of smell/taste above baseline | - | 57 (11.0%) | 392 (43.8%) |
| Total | 7696 | 522 | 895 |

### Association between Covid-19 risk factors and antibody titres

We also considered the relationship between exposure risk factors and antibody titres. Figure 4A shows an ideal assay where low readings are associated with low probability of Covid-19 and increases in assay readings at the diagnostic threshold result in a rapid switch to a high probability of Covid-19. This is analogous to the distributions of assay readings in negative and positive individuals being separate with the diagnostic threshold between them. Figure 4B shows the actual relationship obtained, plotting the mean probability of Covid-19 at some time against antibody titres, using predictions from a model based on healthcare and community Covid-19 exposures, ethnicity, healthcare role, and specialty described in ^3^. The probability of Covid-19 assigned by the model to those with ELISA readings of 0-1 million was 10.2%, which was similar to those with readings of 2-3 million but rose to 11.5% (t-test p=0.02 vs. 0-1 million) by 4-5 million and 11.9% by 6-7 million (p=0.01). A similar trend in point estimates was seen when considering only the greatest risk factor for Covid-19, household contact with a PCR-confirmed case, although the number of individuals with this exposure was less, limiting certainty (Supplementary Figure S3).

**Figure 4.**
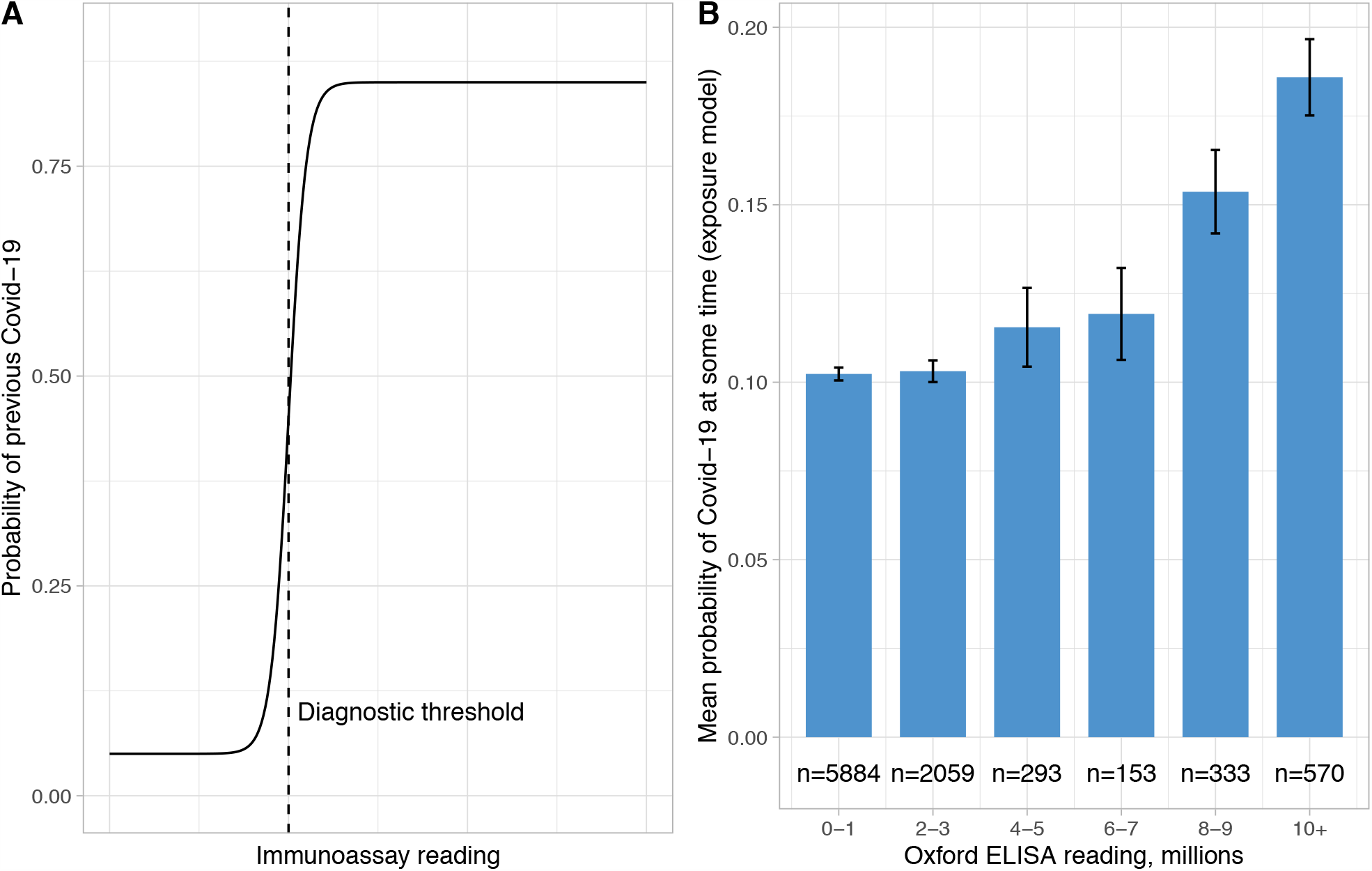
Ideal (panel A) and actual (panel B) relationship between the probability of Covid-19 and antibody titres. The probability of Covid-19 in panel B was generated from a multivariable model containing risk factors including Covid-19 exposures in the community and at work, ethnicity, healthcare worker role and specialty area worked in.^3^

## Discussion

More individuals may have had Covid-19 than are diagnosed using immunoassays calibrated on PCR-positive cases enriched for hospitalised patients. We show, using two different immunoassays that target different SARS-CoV-2 antigens, that intermediate assay results, below diagnostic thresholds for positivity, are associated with increased rates of anosmia, ageusia and other symptoms, as well as being more frequent in individuals with a higher risk of Covid-19. Additionally, we show that around 1 in 10 previously symptomatic and PCR-positive healthcare workers had negative antibody assays when tested between 14 and 62 post a first positive PCR test.

We used self-reported anosmia and ageusia rates across antibody titres to estimate how many Covid-19 cases might be missed by current assay thresholds. Adjusting for rates of anosmia/ageusia in those with high negative (equivocal) antibody titres produces an adjusted test sensitivity for the Oxford ELISA of 90.8% (95%CI 86.1-92.1%) rather than the previously reported 99.1% (97.8-99.7%). Similarly, the adjusted Abbott CMIA sensitivity is 80.9% (77.5-84.3%) compared to 92.7% (90.2-94.8%).^7^ However, we observed several individuals reporting anosmia/ageusia with a positive PCR result but negative antibody titres, including low negative titres (Figure 1). Therefore, as a sensitivity analysis we calculated the adjusted sensitivity assuming an example background rate of anosmia/ageusia of 3%, which lead to adjusted sensitivities for the Oxford ELISA and Abbott CMIA of 75.1% (95%CI 67.6-81.4%) and 70.8% (65.0-79.1%) respectively. This example background is plausible as self-reported rates of anosmia in previous studies vary, e.g. from ∼1 to 5%, with rates of 3% in the ages typical of the healthcare workers studied in one series.^12^ However, these are studies of established objective anosmia, subjective rates may differ, and the question used for our study asked only about new onset loss of smell or taste since 01 February 2020, i.e. excluded established anosmia.

Studies of closed communities or households can also be used to put a lower bound on test sensitivity if universal exposure is considered likely. For example, seroprevalence on the US Navy ship the USS Theodore Roosevelt was 60% following a large outbreak, demonstrating immunoassay sensitivity must be at least this amount (a further 6% were PCR-positive, but antibody negative, possibly reflecting sampling shortly after Covid-19 onset before antibody levels could rise).^9^

Our study suggests it may be appropriate to report an equivocal category of antibody results. This is in contrast to the approach adopted by many manufacturers of reporting with a binary cut-off, for example, of four major commercial platforms compared in a head-to-head evaluation,^7^ only one manufacturer provides an equivocal zone, which is largely intended for guiding repeat testing rather than final reporting. An equivocal category may be particularly informative in individuals with a high pre-test probability of infection where the combined evidence would support previous infection. However, any addition of an equivocal category needs to be balanced against the potential for false-positive equivocal results. In our dataset 387/9292 (4.2%) Oxford ELISA results would be classed as equivocal and 522/9113 (5.7%) Abbott results.

Our study is limited by the lack of a pre-pandemic control group asked the same question regarding new onset loss of taste or smell between February and May of a previous year.

There is therefore uncertainty about the background rate of anosmia/ageusia, and the attributability of this to other respiratory viral causes in this context.^13,14^ Reported anosmia/ageusia may also vary between settings and populations, for example less anosmia is reported in East Asian patients with Covid-19,^15^ however we found similar rates of anosmia/ageusia in staff from Asian ethnic groups (predominantly south and south-east Asian) with Covid-19 to staff identifying as white. The lack of serially collected PCR data for all individuals in our study is another limitation. For example, we cannot say what proportion of those with equivocal antibody responses and anosmia/ageusia would have had a positive PCR test if tested shortly after infection. Similarly, serial antibody tests may have shown rising titres, although 99% of all those reporting anosmia/ageusia and providing a date of symptom onset were tested ≥14 days after symptom onset. Low initial antibody titres may also have waned by the point of testing too. A more general limitation is the absence of a gold standard for retrospective diagnosis of previous Covid-19. Follow up studies are needed to evaluate the extent to which individuals with symptoms or exposures strongly suggestive of Covid-19 such as anosmia or a PCR-confirmed household contact, but negative antibody results, have other evidence of infection, for example from T cell assays. Specific Covid-19 T cell responses have been reported in seronegative individuals who have been exposed to SARS-CoV-2.^16^

The sensitivity of SARS-CoV-2 serology in those with mild symptoms, i.e. the majority of the infected population, is likely to be lower than previously reported, i.e. 90% or less. However, even in the more extreme case that the sensitivity of serology for Covid-19 is as low as 60-70%, population estimates of previous infection in most settings would still be well below those needed for herd immunity, but still sufficiently different to currently understood values to have implications for epidemiological modelling and forecasting. However, it is also important to understand the relationship between antibody titres and neutralisation and correlates of immunity. If low titres in response to mild disease don’t translate into protection this would also have important implications.

For individuals with a high pre-test probability of Covid-19, negative serology does not exclude previous infection, and this may be more common than previously appreciated. Samples from individuals with mild and asymptomatic infection should be included in SARS-CoV-2 immunoassay evaluations in sufficient numbers to assess sensitivity in different populations. Consideration should be given to reporting equivocal results for immunoassays where there is evidence of Covid-19 in some individuals with results below current diagnostic thresholds, and in designing algorithms for reporting that take into account previous symptomatology and pre-test probability.

## Methods

### Setting, participants and immunoassays

Asymptomatic healthcare workers from across 4 teaching hospitals in Oxfordshire, UK, were invited to participate in voluntary staff testing for Covid-19 by nasal and oropharyngeal swab PCR and serological testing. The cohort and associated methods have been previously described in detail,^3^ however the data presented here include additional immunoassay results for some individuals that were not available at the time of the previous publication. Serology for SARS-CoV-2 IgG to nucleocapsid protein was performed using the Abbott Architect i2000 chemiluminescent microparticle immunoassay (CMIA; Abbott, Maidenhead, UK). Samples were also tested by a high-throughput enzyme-linked immunosorbent assay (ELISA) developed at the University of Oxford detecting antibodies to trimeric spike antigen.^7,17,18^

### Analysis

All staff with a sample tested by the Oxford ELISA assay were included in the current analysis. The majority of these samples were also tested using the Abbott CMIA (numbers described in the Results). The first sample tested per individual was included in the analysis for staff tested more than once. Immunoassay titres were compared with the proportion of staff reporting anosmia or ageusia, as well as other symptoms. Self-reported loss of smell or taste was associated with an odds ratio for Covid-19 of 17.7 (95%CI 14.1-22.3) in our earlier study after adjustment for other symptoms.^3^

We also considered the association of different antibody titres with risk factors for Covid-19, including living with a person with PCR-confirmed Covid-19, which had an odds ratio for Covid-19 of 4.63 (95%CI 3.30-6.50) after adjusting for other Covid-19 exposures, role, specialty area worked in and ethnicity.^3^ We also compared antibody titres with the predicted probability of Covid-19 from the same multivariable regression model.

We investigated if age, gender, ethnicity, specialty area worked in or role affected the likelihood of reporting anosmia given an individual was subsequently shown to be SARS-CoV-2 IgG positive. Univariable and multivariable linear regression models were fitted, using natural cubic splines to account for non-linearity of continuous predictors, choosing the best fitting model on backwards selection and number of spline knots using AIC values.

The previously defined and manufacture’s thresholds for confirming detection of anti-SARS-CoV-2 IgG are 8 million and 1.4 for the Oxford ELISA and Abbott CMIA respectively. We used the distribution of the proportion of staff reporting loss of smell/taste within varying antibody titre groups to define a range of titres which we considered “high negative” (equivocal) (see Results, 4.0-7.9 million for the Oxford ELISA and 0.20-1.39 for the Abbott CMIA). We used the proportion of individuals with low negative antibody titres (Oxford ELISA <4 million, Abbott CMIA <0.2) reporting loss of smell or taste to estimate the background rate of anosmia/ageusia unrelated to Covid-19. We subtracted this background proportion (*P*_*baseline*_) from the proportion of anosmia/ageusia in individuals with high negative (potentially equivocal) antibody results (*P*_*eq*_) and the proportion with positive antibody results (*P*_*pos*_) to estimate the overall proportion of anosmia/ageusia attributable to Covid-19. This allowed us to estimate adjusted sensitivity (*S*_*adj*_) of the Oxford and Abbott immunoassays, compared with the previously reported sensitivity (*S*), accounting for the excess of anosmia/ageusia in individuals with elevated but negative antibody results. We denote the number of individuals with positive antibody results *N*_*pos*_ and the number with high negative results *N*_*eq*_ such that:

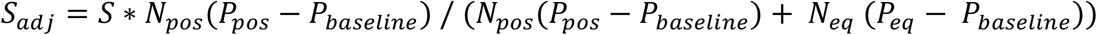

Bootstrapping with 1000 iterations was used to estimate the uncertainty in adjusted sensitivity results, accounting for variation in previously reported sensitivity as well as variation arising from the proportions from the current analysis. We also undertook a sensitivity analysis assuming a proportion of the anosmia/ageusia in those with low negative antibody titres was due to Covid-19.

All analyses were undertaken using R version 3.6.3. Exact binomial confidence intervals are presented for proportions.

### Ethics

All asymptomatic staff data collection and testing were part of enhanced hospital infection prevention and control measures instituted by the UK Department of Health and Social Care. Deidentified data from staff testing were obtained from the Infections in Oxfordshire Research Database (IORD) which has generic Research Ethics Committee, Health Research Authority and Confidentiality Advisory Group approvals (19/SC/0403, 19/CAG/0144).

## Data Availability

The data studied are available from the Infections in Oxfordshire Research Database (https://oxfordbrc.nihr.ac.uk/research-themes-overview/antimicrobial-resistance-and-modernising-microbiology/infections-in-oxfordshire-research-database-iord/), subject to an application meeting the ethical and governance requirements of the Database.

## Acknowledgements

We would like to thank all those who participated in the OUH asymptomatic staff screening programme, and the staff and medical students who ran the programme. This work uses data provided by healthcare workers and collected by the UK’s National Health Service as part of their care and support. We thank all the people of Oxfordshire who contribute to the Infections in Oxfordshire Research Database. Research Database Team: L Butcher, H Boseley, C Crichton, DW Crook, DW Eyre, O Freeman, J Gearing (community), R Harrington, K Jeffery, M Landray, A Pal, TEA Peto, TP Quan, J Robinson (community), J Sellors, B Shine, AS Walker, D Waller. Patient and Public Panel: G Blower, C Mancey, P McLoughlin, B Nichols.

## Declarations

DWE declares lecture fees from Gilead, outside the submitted work. RJC is a founder shareholder and consultant to MIROBio, work outside the submitted work. No other author has a conflict of interest to declare.

## Funding

This study was funded by the UK Government’s Department of Health and Social Care. This work was supported by the National Institute for Health Research Health Protection Research Unit (NIHR HPRU) in Healthcare Associated Infections and Antimicrobial Resistance at Oxford University in partnership with Public Health England (PHE) [grant HPRU-2012-10041] and the NIHR Biomedical Research Centre, Oxford. The views expressed in this publication are those of the authors and not necessarily those of the NHS, the National Institute for Health Research, the Department of Health or Public Health England.

DWE is a Robertson Foundation Fellow and an NIHR Oxford BRC Senior Fellow. SFL is a Wellcome Trust Clinical Research Fellow. DIS is supported by the Medical Research Council (MR/N00065X/1). PCM holds a Wellcome Intermediate Fellowship (110110/Z/15/Z) and an NIHR Oxford BRC Senior Fellow. BDM is supported by the SGC, a registered charity (number 1097737) that receives funds from AbbVie, Bayer Pharma AG, Boehringer Ingelheim, Canada Foundation for Innovation, Eshelman Institute for Innovation, Genome Canada through Ontario Genomics Institute [OGI-055], Innovative Medicines Initiative (EU/EFPIA) [ULTRA-DD grant no. 115766], Janssen, Merck KGaA, Darmstadt, Germany, MSD, Novartis Pharma AG, Pfizer, São Paulo Research Foundation-FAPESP, Takeda, and Wellcome. BDM is also supported by the Kennedy Trust for Rheumatology Research. GS is a Wellcome Trust Senior Investigator and acknowledges funding from the Schmidt Foundation. TMW is a Wellcome Trust Clinical Career Development Fellow (214560/Z/18/Z).

## Contributions

Conceived and designed the study: DWE, TEAP, TMW, KJ, CPC

Secured funding for the study: TMW, KJ

Data collection web application: DWE

Project management: SFL, TMW, DOD, DWE, AH, DE.

Collected the data: DWE, SFL, DOD, NES, PCM, AH, SBH, BDM, SC, TJ, DE.

Supervision: RJC, DWC, DIS, GS, DE, TEAP, CPC, DWE, KJ, TMW.

Curated and analysed the data: DWE, TEAP.

Produced the visualisations: DWE.

Wrote the first draft of the manuscript: DWE.

All authors contributed to editing and revising the manuscript.

## Supplementary figures

**Supplementary Figure S1.**
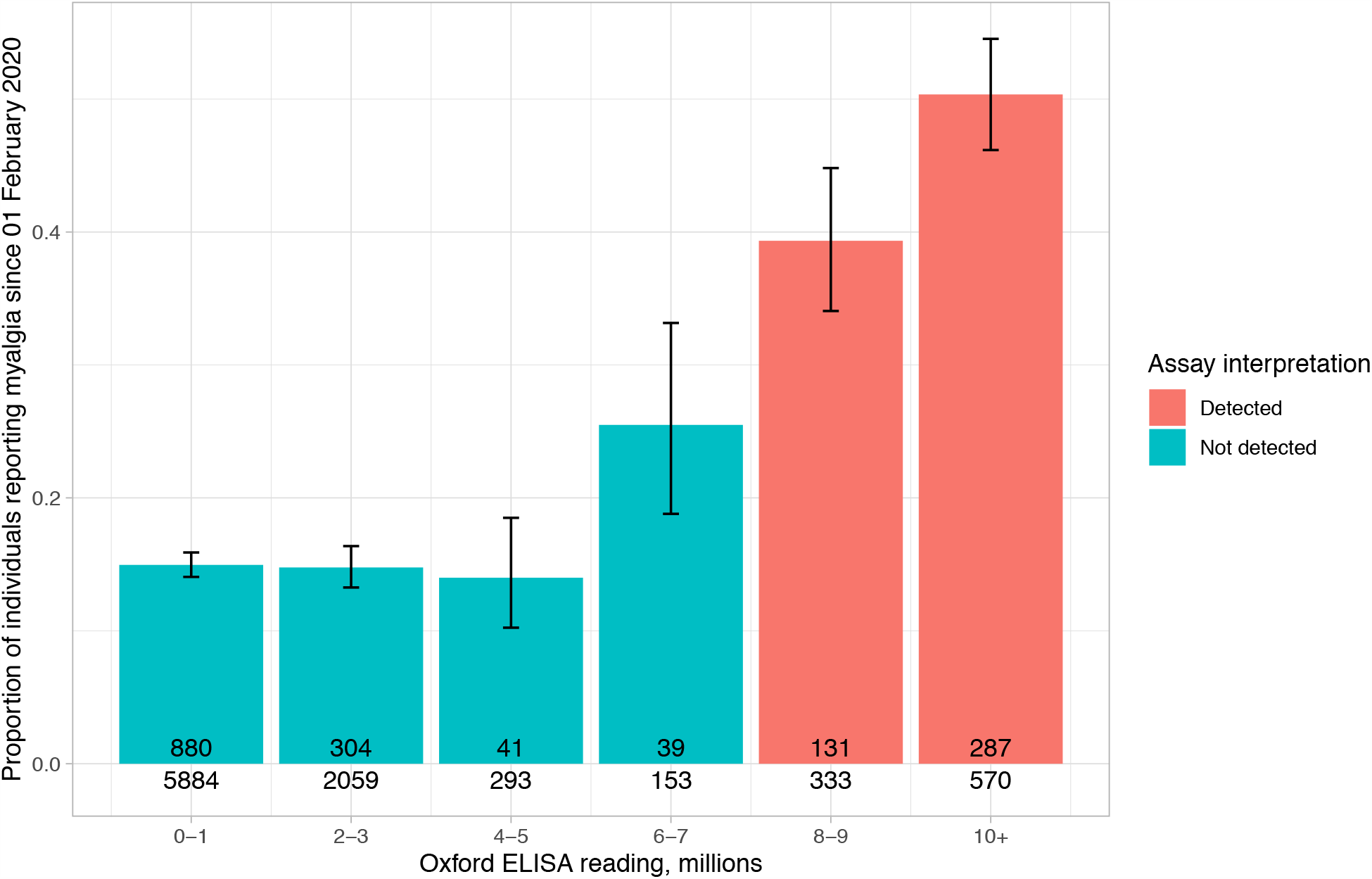
Association of previous myalgia with immunoassay titres. The number of individuals with myalgia is shown in each bar, and the total number of individuals with each antibody titre below the bar. The error bars show 95% confidence intervals. Each value is rounded down, such that for example a value of 1.7 million is within the 0-1 million bar.

**Supplementary Figure S2.**
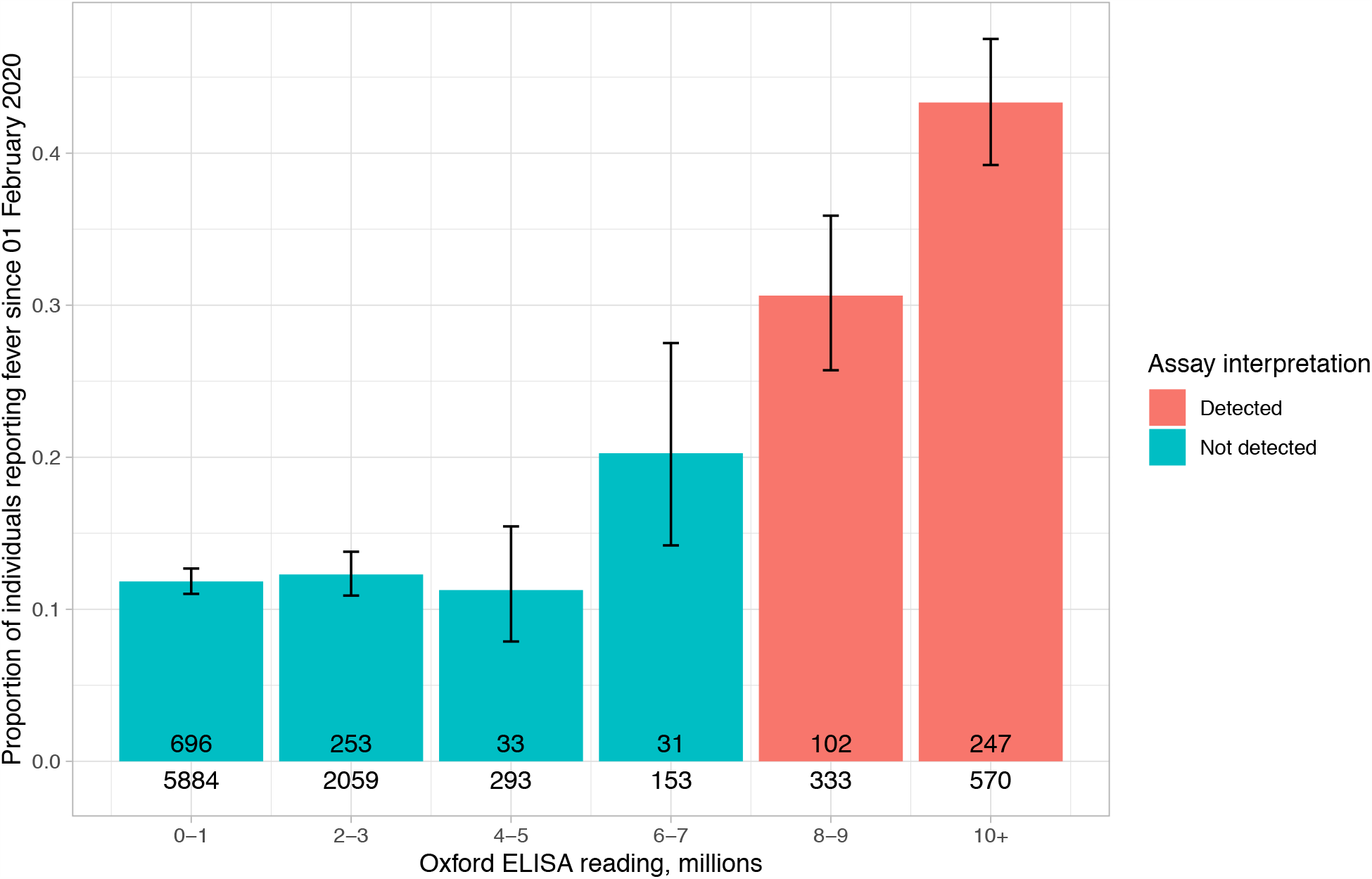
Association of previous fever with immunoassay titres. The number of individuals with previous fever is shown in each bar, and the total number of individuals with each antibody titre below the bar. The error bars show 95% confidence intervals. Each value is rounded down, such that for example a value of 1.7 million is within the 0-1 million bar.

**Supplementary Figure S3.**
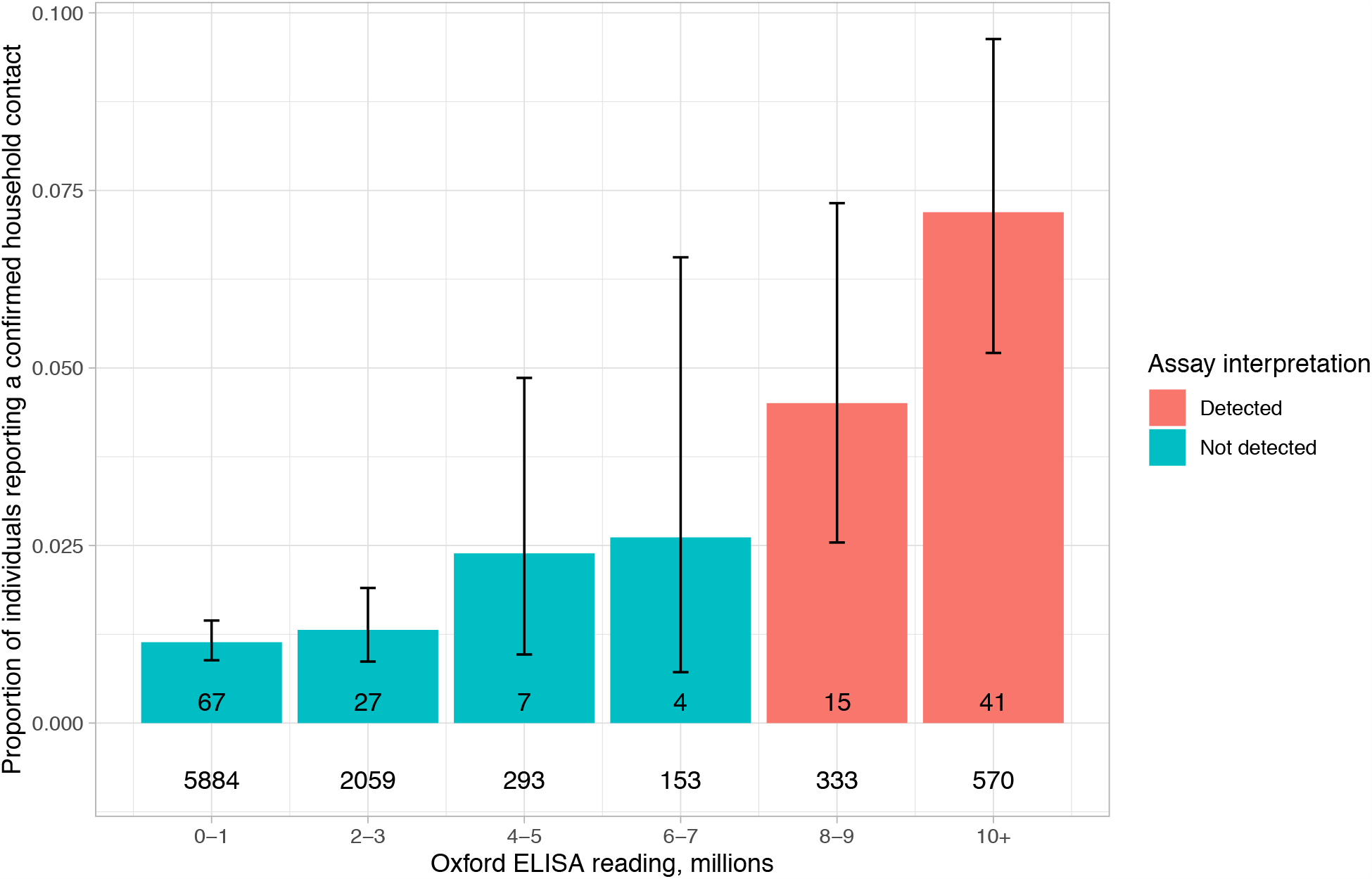
Association of a PCR-confirmed household contact with immunoassay titres. The number of individuals with a PCR-confirmed household contact is shown in each bar, and the total number of individuals with each antibody titre below the bar. The error bars show 95% confidence intervals. Each value is rounded down, such that for example a value of 1.7 million is within the 0-1 million bar.

